# Artificial Intelligence-Enabled Electrocardiogram for Elevated Left Ventricular Filling Pressure

**DOI:** 10.1101/2025.10.03.25337299

**Authors:** Jaehyun Lim, Min Sung Lee, Jung Ho Suh, Sora Kang, Hak Seung Lee, Jong-Hwan Jang, Jeong Min Son, Joon-Myoung Kwon, Yong-Jin Kim, Kyung-Hee Kim, Seung-Pyo Lee

## Abstract

**Background:** Left ventricular filling pressure (LVFP) is associated with heart failure symptoms, a key prognostic marker, and a therapeutic target, but is difficult to measure non-invasively. We aimed to develop and validate a deep learning-based artificial intelligence (AI) model using a standard 12-lead electrocardiogram (ECG) to detect elevated LVFP and assess its prognostic value.

**Methods:** We trained an AI model to detect increased LVFP. Septal E/e’ >15 on Doppler echocardiography was used to define increased LVFP and guide AI-ECG model training. The model was built upon a foundation model trained with >1 million multi-ethnic ECGs and fine-tuned through a development cohort of 225737 ECGs and 115982 echocardiogram data from 92775 unique patients from two tertiary hospitals. The model performance was assessed in a separate internal population from the development cohort (n=9278) and an independent external cohort from another tertiary hospital (n=17926). The prognostic significance of the AI-ECG output value was evaluated via survival analyses using the internal and external hospital cohorts, as well as the UK Biobank (n=43347).

**Results:** The AI-ECG model detected increased LVFP with an area under the curve of 0·868 (95% confidence interval [CI] 0·859–0·877) and 0·850 (95% CI 0·841–0·858) in the internal and external test cohorts, respectively. The model output was an independent predictor of mortality in all three cohorts (adjusted hazard ratio per 10-point increment: internal 1.31 [95% CI 1·23–1·38]; external 1·32 [95% CI 1·28–1·35]; UK Biobank 1.16 [95% CI 1·07–1·26]; all p<0·001). Its prognostic capability was comparable or superior to traditional echocardiographic parameters, particularly in patients with comorbidities.

**Conclusions:** The AI-ECG may enable identification of patients with increased LVFP and provide powerful prognostic information. Further prospective studies are warranted to evaluate its clinical utility.

**CLINICAL PERSPECTIVE:** *What Is New?:*

- By using a specific, broadly applicable echocardiographic marker, E/e’ > 15 as the training target, our model circumvents the well-documented problems of indeterminate classifications and the exclusion of patients with atrial fibrillation, that have constrained previous models.
- The most significant added value is the extensive external validation. We built our model upon a state-of-the-art, multi-ethnic foundation model pre-trained on >1 million ECGs, and demonstrated the model’s consistent high performance not only in an internal cohort but also in two independent, racially and geographically distinct external cohorts. This robust external validation directly confronts the critical challenge of generalizability.

*What Are the Clinical Implications?:*

- The AI-ECG output value provides independent and meaningful prognostic information, with performance comparable or numerically superior to established traditional echocardiographic parameters. This was particularly evident in patients with comorbidities, where the role of traditional echocardiographic markers is often limited.
- The AI-ECG may enable both population-level screening and enhance longitudinal management, offering an opportunity to identify at-risk individuals earlier and implement preventive strategies.

## INTRODUCTION

Elevated left ventricular filling pressure (LVFP), a consequence of left ventricular diastolic dysfunction (LVDD), is associated with heart failure symptoms, and a key prognostic marker and therapeutic target. ^1–6^ However, direct measurement of LVFP via invasive methods, the current gold standard, is impractical for routine screening or therapeutic guidance.

Accordingly, there has been ongoing research to detect increased LVFP noninvaisvely. ^1,5,7^ Several echocardiographic parameters—such as peak early (E) and late (A) mitral inflow velocities, early diastolic mitral annulus velocity (e’), the E/e’ ratio, peak velocity of tricuspid regurgitation by continuous-wave Doppler, and left atrial volume index—have been suggested, ^7,8^ with the latest guidelines integrating these parameters into a diagnostic algorithm. ^1^ Nonetheless, the performance of this algorithm and its individual components has been modest. ^7,9^ Furthermore, each parameter has inherent limitations, particularly notable with the latest guideline, which leaves many patients in an indeterminate category and is inapplicable to patients with atrial fibrillation/flutter (AF/AFL). ^1,7,10^

Artificial intelligence (AI) has provided novel solutions in medicine, particularly by leveraging conventional yet widely-used techniques, such as the electrocardiogram (ECG). ^11–15^ As the ECG encodes the vector summation of electrical signals including myocardial contraction and relaxation, deep neural network-based processing of the ECG has demonstrated potential in unraveling meaningful patterns, suggesting AI-enhanced ECG as a valuable screening and diagnostic tool in various cardiac diseases. ^11–15^

We hypothesized that AI-driven ECG analysis could identify elevations in LVFP. This study aimed to develop and validate an AI model, trained using septal E/e’ >15 as a reference, that performs reliably across diverse ethnicities and populations as well as cardiac rhythms, including atrial arrhythmias. ^8^ The model performance and its prognostic significance were validated in an internal cohort and two independent external cohorts from racially and geographically diverse populations.

## METHODS

### Study design, datasets, and population

This was a retrospective multicenter study involving three tertiary hospitals in South Korea and the UK Biobank. The model was developed using data from adult patients at Bucheon Sejong Hospital (Hospital A) and Incheon Sejong Hospital (Hospital B) who underwent transthoracic echocardiography and a 12-lead ECG within a 14-day window between September 2016 and October 2023. The model was tested both internally and externally, with external testing using data from Seoul National University Hospital (Hospital C). Across all hospital cohorts, patients with a history of mitral valve repair or replacement, a left ventricular assist device, constrictive pericarditis, or missing septal E/e’ were excluded. Raw ECG data were extracted in XML format at a 500Hz sampling rate.

Given that increases in LVFP predict survival, ^2,4,5^ we conducted survival analyses using both the hospital-based mortality data (from Hospitals A, B, and C, for test datasets only) and the UK Biobank data (**Figure 1**). Mortality data were obtained from death statistics provided by Statistics Korea. In the UK Biobank data, resting 10-second ECG recorded at 500Hz was matched with clinical information collected between May 2014 and July 2022.

**Figure 1.**
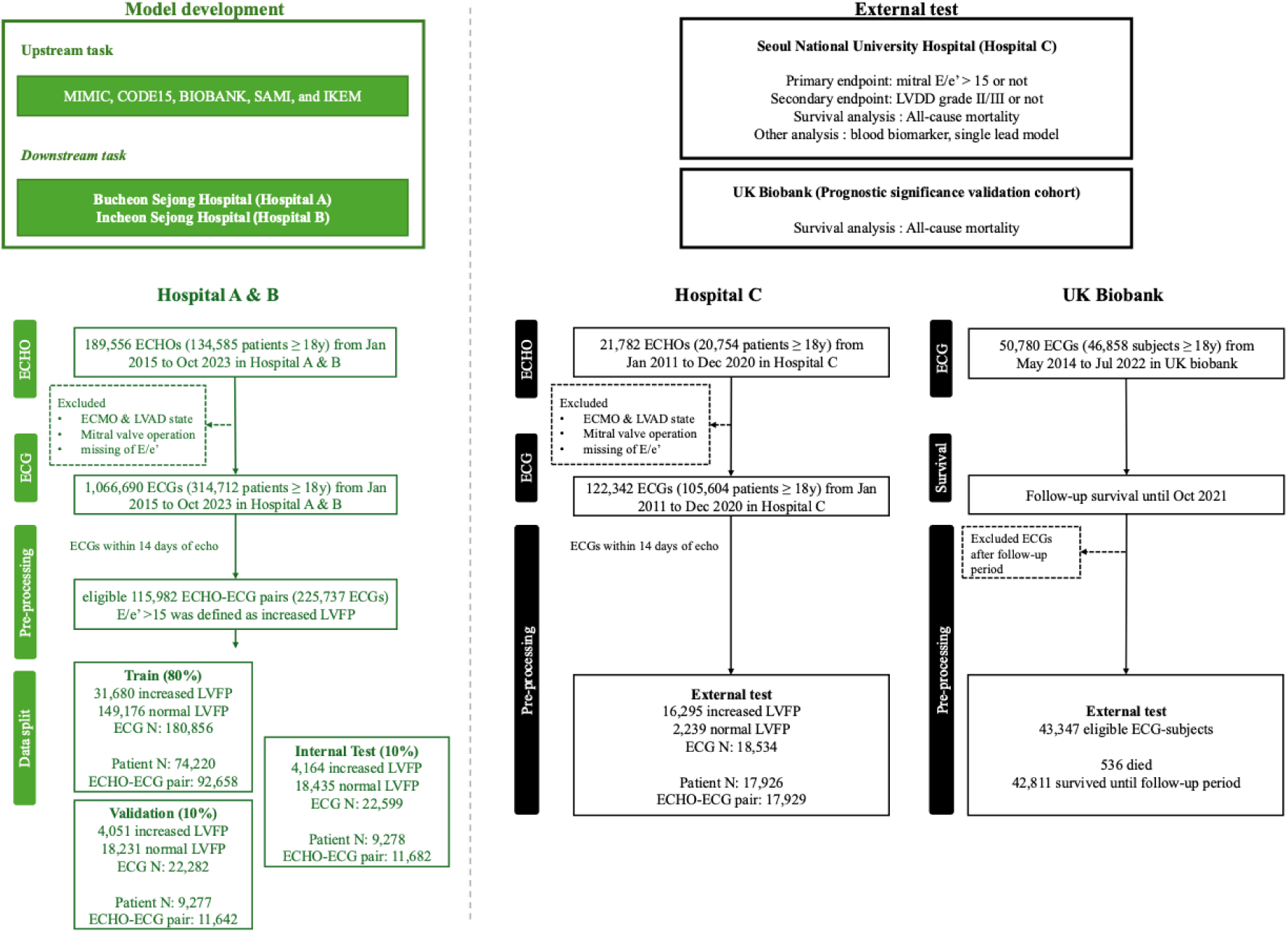
Study workflow. The study outlines the AiTiALVDD model development process, comprising upstream and downstream tasks using large-scale ECG and echocardiogram datasets. External validation was conducted at Seoul National University Hospital and UK Biobank to assess primary, secondary, survival, and exploratory outcomes.

This study was approved by the Institutional Review Boards at each participating hospital (No.: Hospital A, 2024-09-003; B, 2024-09-001; C, H-2408-112-1562), and informed consent was waived. The UK Biobank application (Ref: 89372) was approved with an appropriate Material Transfer Agreement.

### AI-ECG model development

Our model, AiTiALVDD v0.90.00, was built upon a pre-trained foundation model that serves as a generalizable learning framework trained on a large-scale ECG dataset before being fine-tuned for specific clinical tasks. The foundation model was initially trained using 1291868 ECG samples from 442736 unique patients from five international public repositories. The foundation model was subsequently fine-tuned using labeled ECG data from Hospitals A and B to detect increased LVFP. We chose a mitral septal E/e’ ratio >15 as an estimator of increased LVFP. ^8^ Our rationale for using this criterion included its mathematical background linking it to LVFP, its diagnostic accuracy comparable to the latest 2016 ASE/EACVI guideline when validated against invasively measured LVFP, and its broad applicability across cardiac rhythms. ^7,9^ The final model generates a continuous output, an AI-ECG score, between 0–100. A 1-lead version using lead I was also derived from the 12-lead model. Further model specifications are detailed in the **eText 1** and **eFigures 1, 2**.

### Endpoints

The primary endpoint was the ability of the AI-ECG model detecting increased LVFP. The secondary endpoint was the identification of grade II/III LVDD according to the latest guideline. ^1^ Patients with indeterminate LVDD status were excluded from the secondary analysis.

### Survival analysis

We conducted survival analyses using this value in the internal test cohort (Hospital A and B), the external test cohort (Hospital C), and the prognostic significance validation cohort, the UK Biobank (**Figure 1**). Cox proportional hazards models were used to evaluate the prognostic value of the AI-ECG scores. The prognostic discrimination of the AI-ECG score, E/e’ ratio, and LVDD grade for all-cause mortality was compared using C-index.

Subgroup analyses were conducted according to age, sex, comorbidities, electrocardiographic and echocardiographic features (left ventricular ejection fraction [LVEF], mitral regurgitation).

For an exploratory analysis, the AI-ECG score was compared with N-terminal prohormone of brain natriuretic peptide (NT-proBNP) levels. Patients with NT-proBNP levels measured within two days of the echocardiography were included in this analysis.

### Statistical Analysis

Demographic characteristics were compared using Student’s t-test, Mann-Whitney U-test, chi-square test, or Fisher’s exact test, as appropriate. The area under the receiver operating characteristic curve (AUROC), area under the precision-recall curve (AUPRC), sensitivity, specificity, positive predictive value (PPV), and negative predictive value (NPV) with corresponding 95% confidence intervals (CI) were calculated using bootstrap resampling and averaging methods. The optimal threshold for binary classification was selected at the Youden index. The DeLong test was employed to compare AUROCs between models. ^16^ Correlations between two parameters were assessed using the Pearson correlation coefficient.

A multivariable Cox-proportional hazards model was employed to calculate adjusted hazard ratios (aHRs) and 95% CIs, with adjustment for age, sex, hypertension, diabetes mellitus, ischemic heart disease, heart failure, stroke, and CKD. For all Cox regression models, the AI-ECG score was analyzed per 10-point increment. Similarly, age was also analyzed per 10-year increment.

We used R software v4.4.1 and Python v3.9.7, and a two-sided p <.05 was considered statistically significant. This study was reported following the STARD (Standards for Reporting of Diagnostic Accuracy Studies) guidelines.

## RESULTS

### Baseline characteristics: development cohort

The development cohort (Hospitals A, B) included 225737 ECGs and 115982 echocardiograms from 92775 unique patients (**Figure 1** and **Table 1**). Among these, 15130 echocardiogram-ECG pairs (13.0%) had increased LVFP. Patients with increased LVFP were older, more likely to be women, had a higher burden of comorbidities, a larger left atrium, and higher NT-proBNP levels than those with normal LVFP (**Table 1**).

**Table 1.**
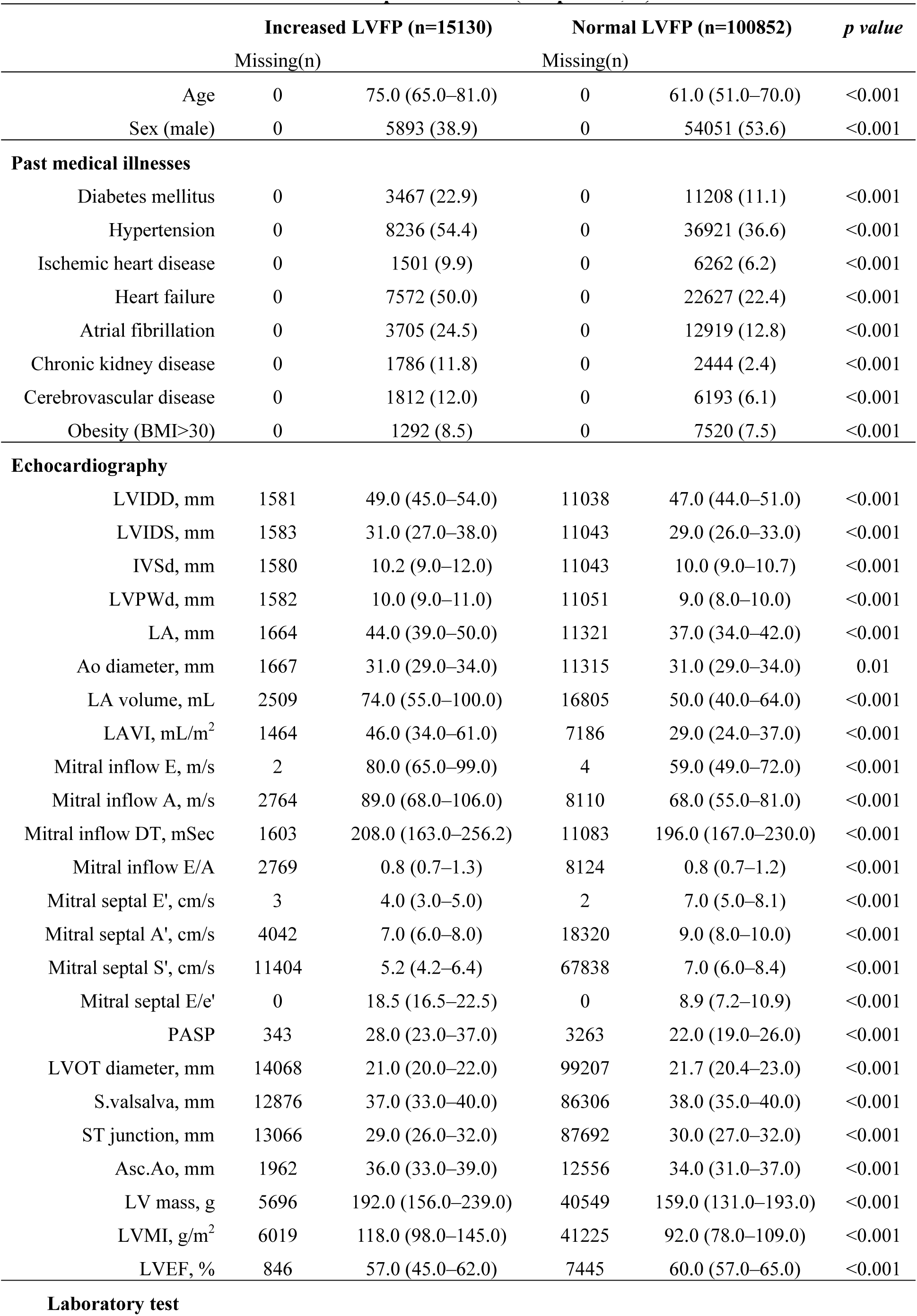
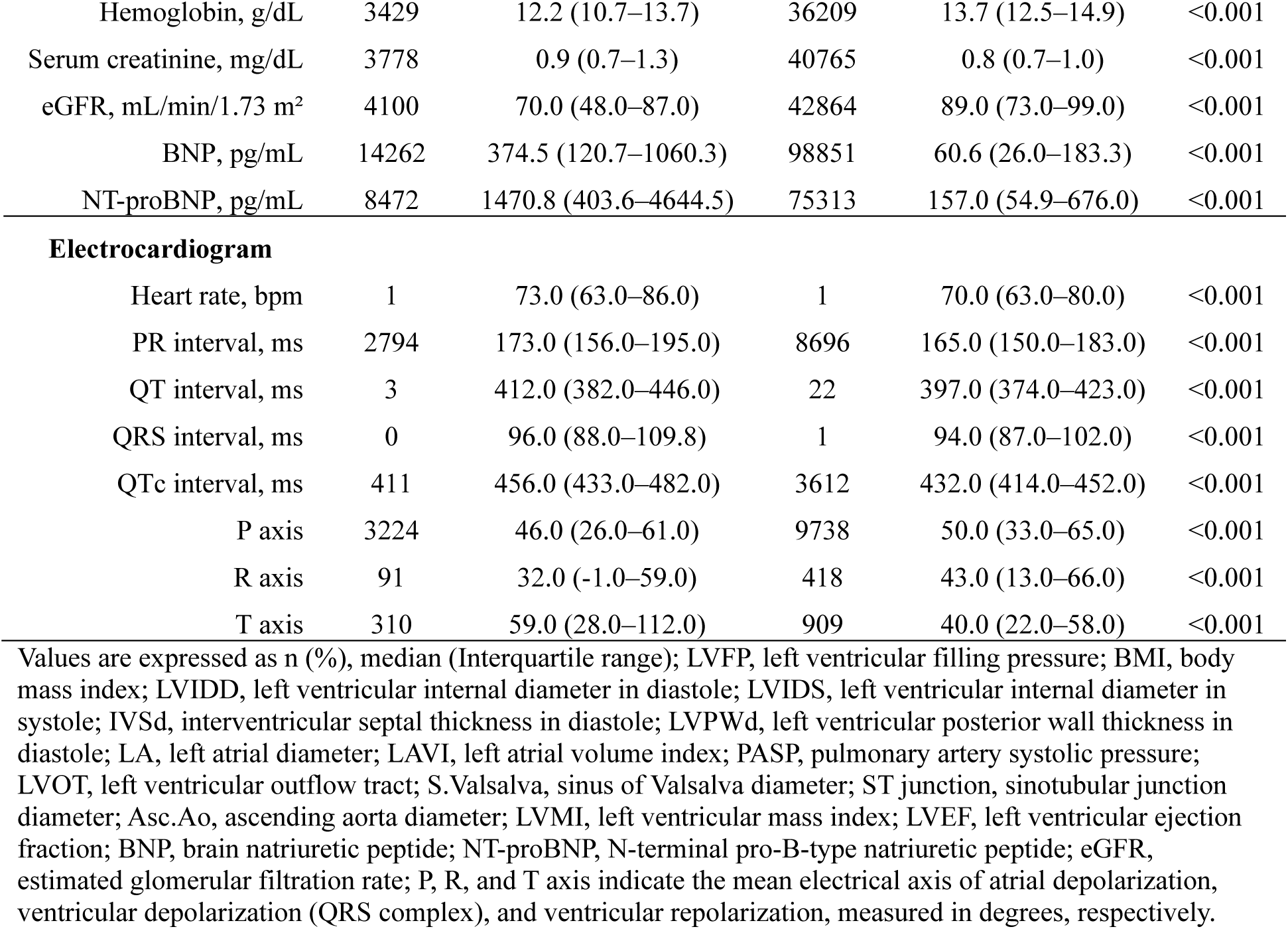
Baseline characteristics: Development cohort (Hospital A, B)

|  | Increased LVFP (n=15130) |  | Normal LVFP (n=100852) |  | *p value* |
| --- | --- | --- | --- | --- | --- |
|  | Missing(n) |  | Missing(n) |  |  |
| Age | 0 | 75.0 (65.0–81.0) | 0 | 61.0 (51.0–70.0) | <0.001 |
| Sex (male) | 0 | 5893 (38.9) | 0 | 54051 (53.6) | <0.001 |
| **Past medical illnesses** |  |  |  |  |  |
| Diabetes mellitus | 0 | 3467 (22.9) | 0 | 11208 (11.1) | <0.001 |
| Hypertension | 0 | 8236 (54.4) | 0 | 36921 (36.6) | <0.001 |
| Ischemic heart disease | 0 | 1501 (9.9) | 0 | 6262 (6.2) | <0.001 |
| Heart failure | 0 | 7572 (50.0) | 0 | 22627 (22.4) | <0.001 |
| Atrial fibrillation | 0 | 3705 (24.5) | 0 | 12919 (12.8) | <0.001 |
| Chronic kidney disease | 0 | 1786 (11.8) | 0 | 2444 (2.4) | <0.001 |
| Cerebrovascular disease | 0 | 1812 (12.0) | 0 | 6193 (6.1) | <0.001 |
| Obesity (BMI>30) | 0 | 1292 (8.5) | 0 | 7520 (7.5) | <0.001 |
| **Echocardiography** |  |  |  |  |  |
| LVIDD, mm | 1581 | 49.0 (45.0–54.0) | 11038 | 47.0 (44.0–51.0) | <0.001 |
| LVIDS, mm | 1583 | 31.0 (27.0–38.0) | 11043 | 29.0 (26.0–33.0) | <0.001 |
| IVSd, mm | 1580 | 10.2 (9.0–12.0) | 11043 | 10.0 (9.0–10.7) | <0.001 |
| LVPWd, mm | 1582 | 10.0 (9.0–11.0) | 11051 | 9.0 (8.0–10.0) | <0.001 |
| LA, mm | 1664 | 44.0 (39.0–50.0) | 11321 | 37.0 (34.0–42.0) | <0.001 |
| Ao diameter, mm | 1667 | 31.0 (29.0–34.0) | 11315 | 31.0 (29.0–34.0) | 0.01 |
| LA volume, mL | 2509 | 74.0 (55.0–100.0) | 16805 | 50.0 (40.0–64.0) | <0.001 |
| LAVI, mL/m<sup>2</sup> | 1464 | 46.0 (34.0–61.0) | 7186 | 29.0 (24.0–37.0) | <0.001 |
| Mitral inflow E, m/s | 2 | 80.0 (65.0–99.0) | 4 | 59.0 (49.0–72.0) | <0.001 |
| Mitral inflow A, m/s | 2764 | 89.0 (68.0–106.0) | 8110 | 68.0 (55.0–81.0) | <0.001 |
| Mitral inflow DT, mSec | 1603 | 208.0 (163.0–256.2) | 11083 | 196.0 (167.0–230.0) | <0.001 |
| Mitral inflow E/A | 2769 | 0.8 (0.7–1.3) | 8124 | 0.8 (0.7–1.2) | <0.001 |
| Mitral septal E', cm/s | 3 | 4.0 (3.0–5.0) | 2 | 7.0 (5.0–8.1) | <0.001 |
| Mitral septal A', cm/s | 4042 | 7.0 (6.0–8.0) | 18320 | 9.0 (8.0–10.0) | <0.001 |
| Mitral septal S', cm/s | 11404 | 5.2 (4.2–6.4) | 67838 | 7.0 (6.0–8.4) | <0.001 |
| Mitral septal E/e' | 0 | 18.5 (16.5–22.5) | 0 | 8.9 (7.2–10.9) | <0.001 |
| PASP | 343 | 28.0 (23.0–37.0) | 3263 | 22.0 (19.0–26.0) | <0.001 |
| LVOT diameter, mm | 14068 | 21.0 (20.0–22.0) | 99207 | 21.7 (20.4–23.0) | <0.001 |
| S.valsalva, mm | 12876 | 37.0 (33.0–40.0) | 86306 | 38.0 (35.0–40.0) | <0.001 |
| ST junction, mm | 13066 | 29.0 (26.0–32.0) | 87692 | 30.0 (27.0–32.0) | <0.001 |
| Asc.Ao, mm | 1962 | 36.0 (33.0–39.0) | 12556 | 34.0 (31.0–37.0) | <0.001 |
| LV mass, g | 5696 | 192.0 (156.0–239.0) | 40549 | 159.0 (131.0–193.0) | <0.001 |
| LVMI, g/m<sup>2</sup> | 6019 | 118.0 (98.0–145.0) | 41225 | 92.0 (78.0–109.0) | <0.001 |
| LVEF, % | 846 | 57.0 (45.0–62.0) | 7445 | 60.0 (57.0–65.0) | <0.001 |
| **Laboratory test** |  |  |  |  |  |
| Hemoglobin, g/dL | 3429 | 12.2 (10.7–13.7) | 36209 | 13.7 (12.5–14.9) | <0.001 |
| Serum creatinine, mg/dL | 3778 | 0.9 (0.7–1.3) | 40765 | 0.8 (0.7–1.0) | <0.001 |
| eGFR, mL/min/1.73 m<sup>2</sup> | 4100 | 70.0 (48.0–87.0) | 42864 | 89.0 (73.0–99.0) | <0.001 |
| BNP, pg/mL | 14262 | 374.5 (120.7–1060.3) | 98851 | 60.6 (26.0–183.3) | <0.001 |
| NT-proBNP, pg/mL | 8472 | 1470.8 (403.6–4644.5) | 75313 | 157.0 (54.9–676.0) | <0.001 |
| **Electrocardiogram** |  |  |  |  |  |
| Heart rate, bpm | 1 | 73.0 (63.0–86.0) | 1 | 70.0 (63.0–80.0) | <0.001 |
| PR interval, ms | 2794 | 173.0 (156.0–195.0) | 8696 | 165.0 (150.0–183.0) | <0.001 |
| QT interval, ms | 3 | 412.0 (382.0–446.0) | 22 | 397.0 (374.0–423.0) | <0.001 |
| QRS interval, ms | 0 | 96.0 (88.0–109.8) | 1 | 94.0 (87.0–102.0) | <0.001 |
| QTc interval, ms | 411 | 456.0 (433.0–482.0) | 3612 | 432.0 (414.0–452.0) | <0.001 |
| P axis | 3224 | 46.0 (26.0–61.0) | 9738 | 50.0 (33.0–65.0) | <0.001 |
| R axis | 91 | 32.0 (-1.0–59.0) | 418 | 43.0 (13.0–66.0) | <0.001 |
| T axis | 310 | 59.0 (28.0–112.0) | 909 | 40.0 (22.0–58.0) | <0.001 |

Values are expressed as n (%), median (Interquartile range); LVFP, left ventricular filling pressure; BMI, body mass index; LVIDD, left ventricular internal diameter in diastole; LVIDS, left ventricular internal diameter in systole; IVSd, interventricular septal thickness in diastole; LVPWd, left ventricular posterior wall thickness in diastole; LA, left atrial diameter; LAVI, left atrial volume index; PASP, pulmonary artery systolic pressure; LVOT, left ventricular outflow tract; S.Valsalva, sinus of Valsalva diameter; ST junction, sinotubular junction diameter; Asc.Ao, ascending aorta diameter; LVMI, left ventricular mass index; LVEF, left ventricular ejection fraction; BNP, brain natriuretic peptide; NT-proBNP, N-terminal pro-B-type natriuretic peptide; eGFR, estimated glomerular filtration rate; P, R, and T axis indicate the mean electrical axis of atrial depolarization, ventricular depolarization (QRS complex), and ventricular repolarization, measured in degrees, respectively.

### Baseline characteristics: external cohort and prognostic significance validation cohort

The external test cohort from Hospital C included 17929 echocardiogram-ECG pairs from 17926 patients, of which 2024 pairs (11.3%) were classified as having increased LVFP (**Figure 1**). The clinical characteristics of these patients were similar to those in the development cohort (**Table 2**). The prognostic significance validation cohort, the UK Biobank, included 43,347 individuals. Baseline characteristics of this cohort are detailed in **eTable 2**.

**Table 2.** Primary and secondary endpoints: AI model discrimination performance of the increased LVFP or LVDD grade (II or III)

|  | **AUROC**<br>**(95% CI)** | **AUPRC**<br>**(95% CI)** | **Sensitivity**<br>**(95% CI)** | **Specificity**<br>**(95% CI)** | **PPV**<br>**(95% CI)** | **NPV**<br>**(95% CI)** | **cutoff** |
| --- | --- | --- | --- | --- | --- | --- | --- |
| **Internal test set** |  |  |  |  |  |  |  |
| **Primary endpoint (n=11682)** | 0.868<br>(0.859–0.877) | 0.540<br>(0.511–0.569) | 0.834<br>(0.816–0.852) | 0.724<br>(0.716–0.733) | 0.311<br>(0.297–0.325) | 0.967<br>(0.963–0.971) | 13.3 |
| **Secondary endpoint (n=10096)** | 0.901<br>(0.892–0.910) | 0.584<br>(0.553–0.614) | 0.856<br>(0.832–0.879) | 0.767<br>(0.758–0.775) | 0.279<br>(0.262–0.294)) | 0.981<br>(0.977–0.984) |  |
| **External test set** |  |  |  |  |  |  |  |
| **Primary endpoint (n=17929)** | 0.850<br>(0.841–0.858) | 0.458<br>(0.435–0.479) | 0.784<br>(0.767–0.801) | 0.749<br>(0.743–0.756) | 0.285<br>(0.273–0.297) | 0.965<br>(0.962–0.968) | 13.3 |
| **Secondary endpoint (n=13301)** | 0.941<br>(0.931–0.951) | 0.487<br>(0.439–0.535) | 0.918<br>(0.891–0.944) | 0.785<br>(0.778–0.791) | 0.116<br>(0.106–0.126) | 0.997<br>(0.996–0.998) |  |

AI, artificial intelligence; LVFP, left ventricular filling pressure; LVDD, left ventricular diastolic dysfunction; CI, confidence interval; AUROC, area under the receiver operating characteristic curve; AUPRC, area under the precision-recall curve; PPV, positive predictive value; NPV, negative predictive value; The pre-specified cutoff (13.3) was determined by Younden's J statistics during model development

### Distributions of AL-ECG model output, the AI-ECG score

In the internal test cohort, the increased LVFP group had a median AI-ECG score of 34.1 (interquartile range [IQR] 18.1–54.8), whereas the normal LVFP group had a median score of 5.6 (IQR 1.6–14.7). A similar separation was observed in the external cohort (median scores 28.4 vs. 6.0, **Figure 2**). There was a stepwise increase in the AI-ECG scores with higher septal E/e’ groups of ≤8, >8 to ≤15, and >15 (**eFigure 3**), as well as LVDD grades based on the 2016 ASE/EACVI recommendations (**eFigure 4**), in both the internal and external test cohorts. The Pearson correlation coefficients between E/e’ and AI-ECG score were 0.60 and 0.54 in the internal and external test datasets, respectively (both p<.001, **eFigure 5**).

**Figure 2.**
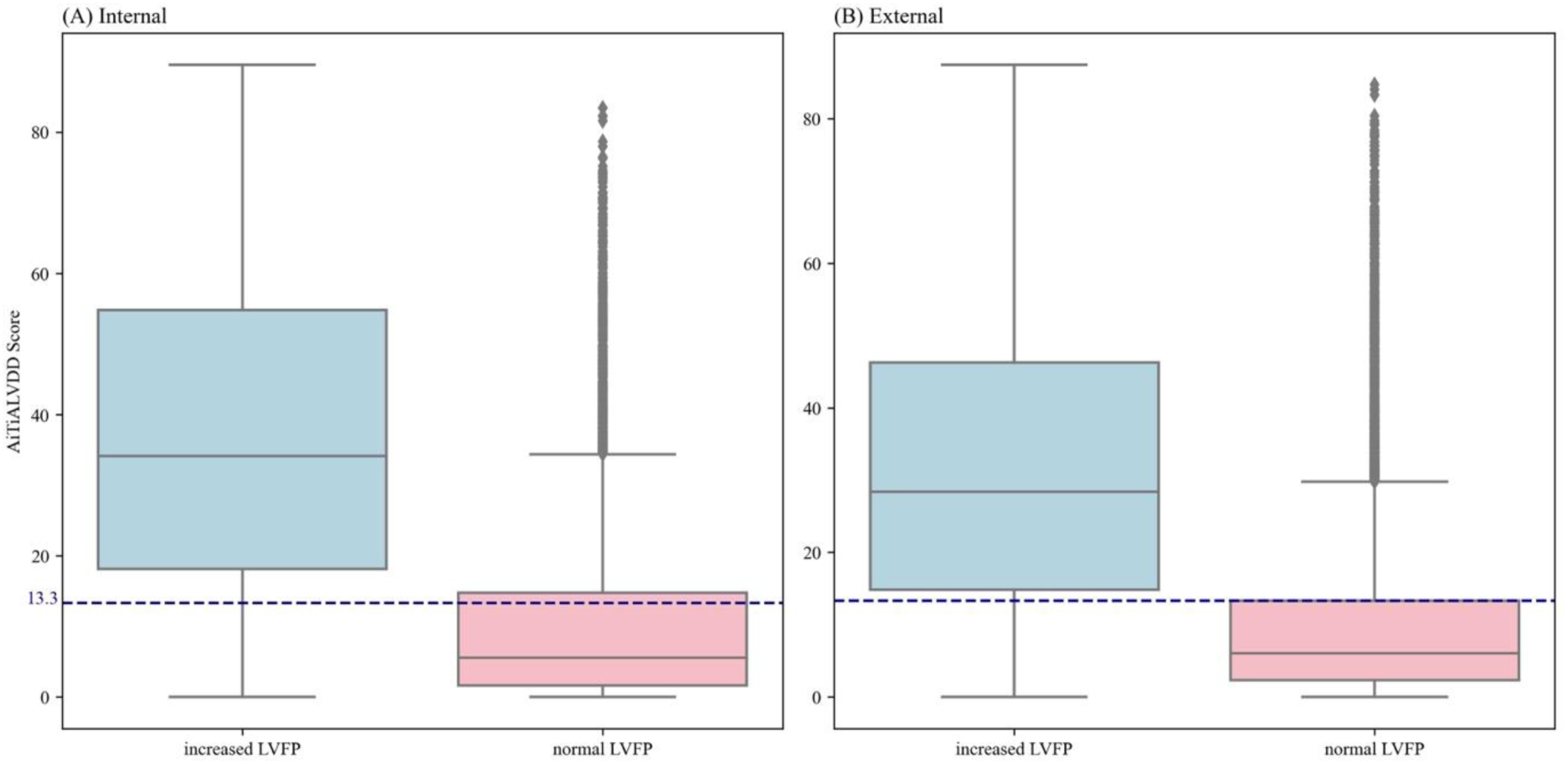
Comparison of the AI-ECG score distribution between increased and normal left ventricular filling pressure groups. Distribution of the AI-ECG scores according to left ventricular filling pressure (LVFP) status. The boxplots illustrate the AI-ECG scores for the increased LVFP group (blue) and the normal LVFP group (pink). In the increased LVFP group (internal test) the mean AI-ECG score was 37.2 ± 22.1, with a median of 34.1 (Interquartile range, 18.1–54.8). In contrast, the normal LVFP group had a mean AI-ECG score of 10.6 ± 13.1 and a median of 5.9 (1.6–14.7). In the increased LVFP group (external test) the mean AI-ECG score was 32.2 ± 20.7, with a median of 28.4 (14.8–46.3). In contrast, the normal LVFP group had a mean AI-ECG score of 10.0 ± 11.7 and a median of 6.0 (2.3–13.3). The cutoff score of 13.3 (indicated by the blue dashed line) was predefined during the model development phase using Younden’s J statistic. Panel (A) represents the internal test, while panel (B) represents the external test.

### Primary and secondary endpoints

For the primary endpoint of detecting increased LVFP, the AI-ECG model achieved an AUROC of 0.868 (95% CI 0.859–0.877) and an AUPRC of 0.540 (95% CI 0.511–0.569) in the internal test cohort. A similar AUROC of 0.850 (95% CI 0.841–0.858) and AUPRC of 0.458 (95% CI 0.435–0.479) were demonstrated in the external test cohort (**Figure 3** and **Table 2**). Sensitivity, specificity, PPV, and NPV were 83.4% (95% CI 81.6–85.2), 72.4% (95% CI 71.6–73.3), 31.1% (95% CI 29.7–32.5), and 96.7% (95% CI 96.3–97.1) in the internal test cohort. The performance was also robust in the external cohort (**Table 2**).

**Figure 3.**
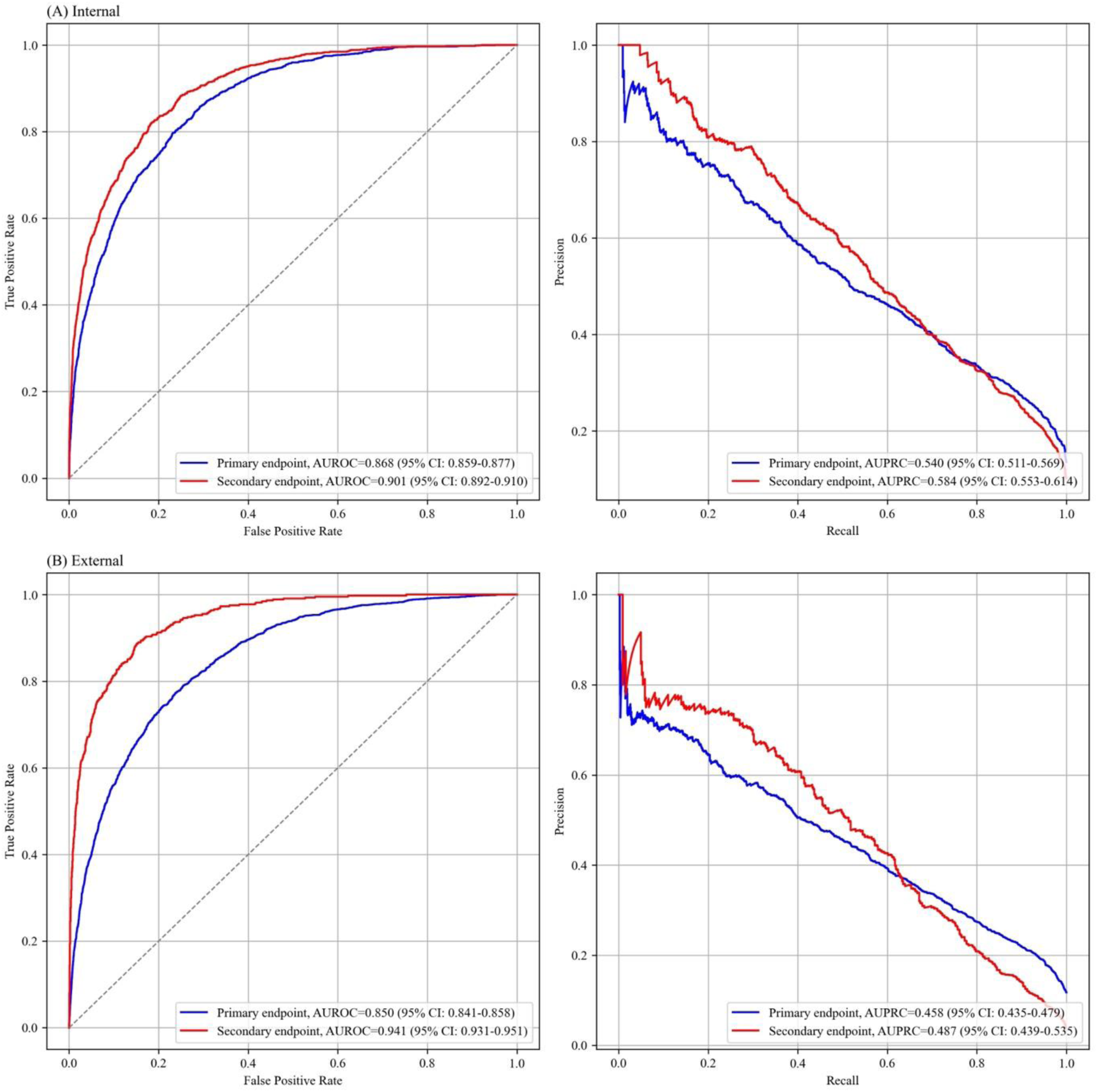
Receiver operating characteristic curve analyses for primary and secondary endpoints. Receiver operating characteristics curves (panel A, internal test; panel B, external test), illustrating the discriminative ability of the AI-ECG model in detecting elevated left ventricular filling pressure, defined by a mitral E/e’ ratio > 15 (primary outcome), and in identifying left ventricular diastolic dysfunction grade II/III (secondary outcome). The primary outcome is depicted in blue, and the secondary outcome is depicted in red. AUROC, area under the receiver operating characteristic curve; AUPRC, area under the precision-recall curve

For the secondary endpoint (grade II/III LVDD), the model demonstrated high AUROCs of 0.901 (95% CI 0.892–0.910) and 0.941 (95% CI 0.931–0.951) in the internal and external test cohorts, respectively (**Figure 3** and **Table 2**).

### Subgroup analysis of primary and secondary endpoints

In the subgroup analyses, the AUROCs were consistently lower in patients with unfavorable clinical characteristics, such as advanced age, comorbidities including hypertension, diabetes mellitus, CKD, ischemic heart disease, systolic dysfunction, or non-sinus rhythms, and AF/AFL, in both the internal and external cohorts (**eTables 3** and **4, eFigure 6**). The performance for discriminating grade II/III LVDD was generally better across subgroups, but the tendency of lower performance in those with unfavorable clinical characteristics was consistently observed (**eTables 5** and **6, eFigure 7**).

### Survival analysis of AI-ECG for increased LVFP

The internal test cohort (Hospitals A and B), external test cohort (Hospital C), and the UK Biobank consisted of 7568, 17922, and 43347 patients, respectively, with median follow-up periods of 2.7 (IQR 1.4–4.0), 5.8 (IQR 3.9–8.0), and 3.4 (IQR 2.5–4.9) years. During follow-up, 361 deaths occurred in the internal test cohort (4.8%), 2837 in the external test cohort (15.8%), and 536 in the UK Biobank (1.2%). Kaplan–Meier analysis showed progressive increases in mortality by AI-ECG score quartiles (**eFigure 8–10**).

After multivariable adjustment, the AI-ECG score remained independently associated with mortality across all cohorts: adjusted hazard ratio (aHR) 1.31 (95% CI 1.23–1.38, p <.001) in the internal test cohort; aHR 1.32 (95% CI 1.28–1.35, p <.001) in the external test cohort; and aHR 1.16 (95% CI 1.07–1.26, p <.001) in the UK Biobank (**eTables 7**–**9**).

Regarding prognostic performance, as measured by the C-index, it was generally favorable for the AI-ECG score compared to echocardiographic LVDD grading and E/e’ ratio (**eTable 10**). In the internal test cohort, the AI-ECG score achieved a C-index of 0.877 (95% CI 0.855–0.898) versus 0.868 (95% CI 0.846–0.890) for LVDD grading and 0.867 (95% CI 0.845-0.889) for E/e’ ratio. In the external test cohort, the C-index for AI-ECG was 0.720 (95% CI 0.708–0.733), compared to 0.711 (95% CI 0.699–0.724) for LVDD grading and 0.709 (95% CI 0.697-0.721) for E/e’ ratio, with no statistically significant difference between AI-ECG and LVDD grading (p=0.305). Notably, the AI-ECG score demonstrated numerically superior prognostic discrimination for all-cause mortality particularly in most subgroups of patients with comorbidities (**eTable 10**).

### Performance comparison between AI-ECG and NT-proBNP for increased LVFP

In our exploratory analysis, the 12-lead AI-ECG outperformed NT-proBNP in both internal and external test cohorts regarding the primary endpoint: AUROCs were 0.840 (95% CI 0.821–0.856) for AI-ECG vs. 0.809 (95% CI 0.788–0.828) for NT-proBNP in the internal test, and 0.835 (95% CI 0.802–0.866) for AI-ECG vs. 0.767 (95% CI 0.730–0.802) for NT-proBNP in the external test cohort (**eTable 11**). Consistent results were demonstrated for secondary outcomes (**eTable 12**). Notably, a single lead model achieved performance comparable to NT-proBNP (**eTables 11** and **12**).

## DISCUSSION

The present study demonstrates that the AI-ECG model can reliably identify increased LVFP, whether defined by an E/e’ >15 or by LVDD grading (grade II/III), and provide robust prognostic information. Importantly, these results were validated in racially and geographically diverse external cohorts, highlighting the model’s generalizability.

Deep neural networks, when trained with sufficiently large datasets, can effectively generalize and accurately discern meaningful patterns even in the presence of considerable noise—even up to 50:1 of noise-to-signal ratios in the presence of a sufficiently large number (≥10,000) of valid samples. ^17^ This has also been demonstrated in medicine. For instance, although diagnostic variability among ophthalmologists in diabetic retinopathy is substantial, AI models trained using these noisy human-labeled datasets have demonstrated performance surpassing that of ophthalmologists. ^18,19^ Our model was pre-trained on over a million ECGs and fine-tuned using over 100,000 ECG–echocardiogram pairs labeled with E/e′. Thus, despite the acknowledged suboptimal nature of E/e’ for defining increased LVFP, this dataset likely provided sufficient volume and variety for the AI model to learn ECG patterns representative of elevated LVFP, resulting in comparable or superior performance in certain contexts to traditional echocardiographic parameters.

Selecting a reference standard for AI model development requires balancing precision with data availability and generalizability. While invasive measurements are the gold standard, their relative scarcity limits dataset size and increases risk of overfitting. Indeed, we, for the first time, have reported an AI-ECG model detecting diastolic heart failure with preserved LVEF based on the 2016 ASE/EACVI recommendations, ^1,13^ with a subsequent study demonstrating similar findings across various EF ranges. ^12^ Despite acceptable AUROCs, it should be noted that our previous study was limited to patients with preserved LVEF, while the latter study lacked rigorous external validation. ^12,13^ Indeed, reproducibility and rigorous external validation of AI models are to be emphasized. ^20^ In addition, as these studies utilized LVDD grading, they inherently excluded AF/AFL, significantly limiting generalizability and clinical applicability given its high prevalence. ^1^ The 2016 algorithm also leaves many patients in an indeterminate category. ^10^ Furthermore, the diagnostic accuracy of this integrated algorithm is only comparable to E/e’ in detecting increased LVFP, particularly in patients with preserved LVEF (see **eTable 13** for comparison). ^9^ Thus, our model trained based on E/e’, given its physiological rationale and broader applicability across cardiac rhythms, offers greater clinical acceptability and generalizability. ^7^

LVFP holds two clinical objectives: therapeutic guidance and prognostication. Regarding prognostication, our study demonstrated that the AI-ECG model provides robust prognostic prediction, often comparable or superior to echocardiographic parameters, particularly in patients with comorbidities. The lower hazard ratio in the UK Biobank data is likely attributable to its lower heart failure prevalence (0.7%) compared to hospital-based cohorts enriched with patients having increased LVFP in the presence of systolic dysfunction. As for therapeutic guidance, guiding therapy based on the accurate assessment of LVFP has significant clinical implications, as demonstrated by the CHAMPION trial where invasive hemodynamic-guided therapy significantly reduced heart failure hospitalizations. ^3^ However, such invasive monitoring is impractical for widespread use. While echocardiography has been also demonstrated to guide therapy, its utility for frequent reassessment is limited by high cost, resource demands, and intra- and inter-observer variability. ^5,21,22^ Harmonizing AI with ECG addresses these limitations, enabling frequent, noninvasive monitoring. Moreover, its simple and rapid nature makes it particularly well-suited for population-based screening. ^23^ Its favorable performance compared to established biomarkers such as NT-proBNP further highlights its potential applicability.

Prior research on using AI-ECG to detect increased LVFP is limited. One study trained a model directly on invasively measured pulmonary capillary wedge pressure (PCWP), and reported modest performance (AUROC 0.79–0.82) in internal and subsequent external validation. ^15,24^ It should be noted that this approach is inherently susceptible to selection bias and lacks generalizability despite its methodological robustness, because only high-risk patients necessitating invasive pulmonary artery catheterization were used to train the model. Nonetheless, this prior work should be appraised since they demonstrated the proof-of-concept that changes in model outputs generally correlate with changes in the PCWP, demonstrating the possibility that AI-ECG may be used to track patients and guide treatment.^24^

Subgroup analyses offer a critical insight into the AI-ECG model. We observed a consistent tendency toward lower AUROC in subgroups with advanced age or comorbidities. However, the reduced performance in these subgroups may not reflect a deficiency in the AI model, but rather the inherent limitations of the echocardiographic reference standard: that is, the E/e’ ratio itself may less accurately represent the true elevation in LVFP in these populations, leading to an artificially lower AUROC. ^25,26^ For instance, the subgroup with LVEF <40% provides compelling insight. Prior invasive catheterization studies reported that 80–95% of patients with systolic dysfunction had increased LVFP. ^2,6,27^ Whereas the AI model classified 85.3% of these patients as having increased LVFP, only 39.7% of these patients were classified as such according to the E/e’ or LVDD grading, strongly suggesting that the AI-ECG approach may more accurately reflect true elevations in LVFP. Similarly, previous literature has suggested a lower E/e’ threshold (e.g., E/e’>11) for increased LVFP in patients with AF/AFL. ^26,28^ Accordingly, the observed low specificity in these patients likely indicates that some of those who were classified as having normal LVFP according to E/e’≤15 might actually have increased LVFP. Ultimately, the observation that our model—trained on E/e’ >15 threshold—aligns better with the expected clinical and pathophysiological status in subgroups where that same threshold is known to be unreliable, suggests the model learned underlying physiological signals beyond the label itself.

### Limitations

Our study has several limitations. First, despite its robust learning capability, training the model solely based on echocardiographic parameters may lack clinical robustness. However, using E/e’ for model training is a strategic compromise, obtaining sufficient data volume and minimizing selection bias at the expense of methodological robustness, ultimately ensuring greater model generalizability. Second, more delicate model training, such as using different thresholds of E/e’ according to age or cardiac rhythm, might have improved performance. Third, the optimal clinical thresholds for guiding therapeutic interventions may vary across individuals and remain to be established. Future research is required to define and validate such clinically relevant thresholds. Lastly, a maximum interval of 14 days between ECG and echocardiography might allow for changes in the patient’s clinical status.

## CONCLUSION

The present study demonstrates that an AI-ECG may identify patients with increased LVFP. The AI model showed robust performance in external validation cohorts derived from geographically and racially different populations. Beyond correlation with echocardiographic parameters indicating increased LVFP, our model provided meaningful prognostic information. Further studies are warranted to evaluate the clinical utility of AI-ECG in screening patients with increased LVFP or guiding interventions in these patients.

## Author Contributions

Jaehyun Lim (Study design, Data analysis, Manuscript drafting and critical revision), Min Sung Lee (Data acquisition, Statistical analysis, Manuscript drafting), Jung Ho Suh (Data curation, Data interpretation, Manuscript revision), Sora Kang (Data curation, Statistical analysis, Manuscript revision), Hak Seung Lee (Data curation, Data interpretation, Critical manuscript revision), Jong-Hwan Jang (Data management, Statistical support, Manuscript review), Jeong Min Son (Data acquisition, Data verification, Manuscript review), Joon-Myoung Kwon (Study conception, Model development, Manuscript drafting and critical revision), Yong-Jin Kim (Clinical data interpretation, Manuscript review), Kyung-Hee Kim (Study design, Study supervision, Data analysis, Manuscript drafting, Review and Approval), and Seung-Pyo Lee (Study design, Study supervision, Data analysis, Manuscript drafting, Review and approval).

## Data Availability

The data used for this study are available upon reasonable request to the corresponding author for research purposes. The AiTiALVDD model used in this study will be made available to external researchers upon request to validate and expand the findings presented in this study.

